# Exploring gendered patterns of interaction within an inter-professional health care network

**DOI:** 10.1101/2020.04.07.20057364

**Authors:** Nelson Aguirre, Peter Carswell, Tim Kenealy

## Abstract

**Background:** Numerous studies have shown gender-based similarity affects interactions in organizational contexts. However, studies in the health care arena have shown contradictory findings.

**Objective:** To explore gender homophily in an inter-professional network comprising doctors and nurses across the primary and secondary care interface in diabetes care.

**Methods:** A Social Network Analysis was conducted with primary and secondary care clinicians responsible for diabetes care in Auckland New Zealand. Three different methods were used to test gender affinity in 40 health professionals (GPs, endocrinologist and specialist nurses). First, a metric analysis of homophily ranking corrected for potential differences in gender proportions was conducted. Homophily ranking is scored between −1 (perfect heterophily) and 1 (perfect homophily). We also examined the ego-net composition and measured the density of interactions among men and women in the network.

**Results:** Gender homophily was close to 0, indicated that network members were likely to interact with males and females without preferences, a result that was confirmed through ego-net composition.

**Conclusions:** This study in diabetes managed care network found little evidence to support the impact of gender homophily on communication exchange. This contrasts with other studies in the health care context. Other influences need to be explored at this context.

## BACKGROUND

Collaboration and communication within organizational and inter-professional networks helps diffusion of information and social influence [1-3]. Inter-professional networks have an instrumental value because they affect outcomes performance, allow better access to critical resources and provide social support [4, 5]. Network interactions are grouped or clustered according to the individual’s characteristics such as race, gender, profession and religion among others [6, 7]. A number of studies have focused on gender as a factor that may influence interactions within networks. Some of the studies have shown that there are differences between men and women in the way they can access the benefits of the social networks, and about motivations to strengthen links. Many health professions are numerically dominated by either men or women [8]. However, in spite of substantial research on how gender may help network development in health care [9-11], there are still overlapping and contradictory results [10, 12, 13].

This study focussed on determining how gender may affect the interactions between health professionals at the primary and secondary care interface. This knowledge is useful for planning and designing inter-professional networks working across and within this interface. A social network approach which seeks to identify social network characteristics and patterns of interaction within a network because the social structure of a network effects individual actions and vice versa [14-16]. Social network analysis can identify the distribution of the social interactions within a network and show how they are clustered [14, 17-19].

## LITERATURE REVIEW

### Homophily within inter-professional networks

The way inter-professional networks are organized and perform is based on the interactions among professionals. Some of the characteristics of the interactions are: the content shared (products or services, information), form (duration and closeness of the interactions), intensity (density and frequency of interactions) [20-22], and the tendency of the individuals to interact with those with similar attributes (homophily) [23, 24].

Recent research has examined the effects of homophily according to individuals’ similarity of attributes such as age [20], ethnicity [21, 22], religion [23, 24], profession [8], knowledge [25] and gender [9, 26, 27]. Homophily within networks may increase perceptions of interpersonal trust by simplifying process of evaluation and facilitating communication [27, 28].

Homophily could be induced by the network structure such as the work place, voluntary organizations, schools or friendship circles. Alternatively, network homophily may derive from personal or psychological preferences of network participants [28]. Those preferences may explain variations in homophily within a network, and suggest mechanism in which homophily change over the time.

### Gender homophily

Gender similarity is a substantial subject in the social network literature [10, 21, 23, 29, 30]. It is defined as the association between social characteristics and interactions between males and females. It constitutes a way in which social life is organised, describing patterns of individuals’ interactions based on their gender characteristics [26, 27, 31].

Researchers have studied gender homophily in social networks such as schools, young unmarried men and women and families among others. Their findings illustrate how women and men are orientated to build ties with similar people. For example men maintain their dominance excluding women from their network and tend to have more gender homophily ties [29], and women have more contacts with neighbours and have more family-oriented relationships than men [26]. Men are likely to obtain information from diverse sources as they are inclined to interact with a large number of weak ties [32], while women prefer “small towns” where everyone knows each other, such that they can receive and provide more support from others, establishing closer relationships [26].

However, the context of human interactions within organizations may introduce several differences. Gender homophily has been studied in different contexts such as communication and technology companies [27], academic scientists [25, 33], entrepreneur founding companies [21, 22] and healthcare [6, 34] among others. There are some contradictory results, as some studies highlight the importance of gendered relationships as a factor in differentiation of network structures and as associated with power [27], while others do not find gender differences or affinities to interact and establish channels of communication [22, 35].

### Gender preferences in healthcare networks

Gender homophily in healthcare is characterized by health professionals’ preferences in gender relationships (i.e. a desire to interact with health professionals of the same gender) [23]. Researchers have studied how interpersonal professional relationships (including gender homophily) may affect diffusion of innovations and the adoption of new practices in healthcare [36-40].

Studies have described gender homophily in healthcare teams, highlighting that male doctors usually interact with male colleagues in seeking for help. Women doctors seek supportive and empathic ties and they are more likely to involve women in their inter-professional networks [8, 34]. Female nurses are more likely to build cross-sex relationships than male doctors [8, 41]. However, induced gender homophily (from numerical availability) is a factor to take an account, as some specialties and health care professions are influenced by sex-segregation. As West and Barron (2005) have outlined nurses in UK are predominantly women and medical specialists in neurology, cardio-thoracic medicine and renal medicine are predominantly male while specialists in dermatology, community health and ophthalmology are predominantly female.

Although gender homophily appears to be a critical factor for network and tie development at the individual level, other studies do not find this association within health care teams [42-44], where other socio-demographic factors such as profession or years of experience are more strongly associated with building ties. Identifying factors that facilitate network development in health care could be a strategy for building successful teams, so that if gender is a characteristic that may affect the way in which health professionals share ideas and knowledge, it should be accounted for in attempts to strengthen the relationships between health professionals at the primary and secondary care interface.

## METHODS

The goal of this paper is to explore gender homophily in an inter-professional network comprising the primary and secondary care interface in diabetes managed care. We hypothesise that the distribution of the interactions is influenced by health professionals’ gender. Interactions or ties were defined as formal and informal communication, advice, seeking help or benefit, sharing of resources, information flows, or some other form of inter- professional exchange between health professionals at the primary and secondary care interface [45, 46].

### Design

A quantitative cross-sectional study conducted in March 2012 was used to collect socio- demographic information and data for social network analysis.

### Setting

Participants were primary care and secondary care doctors and nurses in South Auckland New Zealand.

### Participants

We invited health professionals from primary care and from secondary care who were involved in the care of the same group of diabetic patients. The first group consisted of general practitioners (GPs) and practice nurses (PNs). The second group included endocrinologists and diabetes nurses specialists (DSNs) who work within the secondary care diabetes service. We invited all GPs located in Counties Manukau and were members of the same Primary Health Organisation and the secondary care group were contacted directly at the main referral hospital at the same area.

### Procedures

This study received ethics approval from the New Zealand Ministry of Health’s Northern Regional Health and Disability Ethics Committee NTX/11/EXP/150 dated 19/07/2011. Participants completed a social network questionnaire delivered online using LimeSurvey^®^ software. Reponses were collected electronically. Data were checked, cleaned, and then analysed in SPSS for Windows V.19. The information collected included demographic characteristics, the nature of the relationship (role relationship i.e. GP – specialist) between the respondent with each of his or her colleagues – known as “alters” in the network literature – and the frequency, quality and perceived value of the interactions between health professionals.

For the first method of analysis, four variables were used to calculate homophily (H) using the following formula:

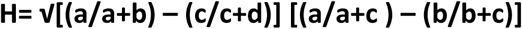

where (a) is the number of ties a person sent to the people of the same gender, (b) is number of ties that an individual sent to the people of the opposite gender, (c) is the number of people of the same gender that the actor could have cited but did not, and (d) is the number of people of the opposite gender the actor could have cited but did not. This calculation effectively corrects for induced homophily, i.e. apparent homophily induced simply by imbalances of gender of potential network-tie partners [29, 47, 48]. Calculated H ranges from −1 to 1. Positive values indicate the tendency of individuals to select actors of the same gender in the network (homophily). A zero value indicates a balance between selecting male or female and negative values indicates the tendency to elect actors of the opposite gender (heterophily).

In the second analysis, Ego-net homophily was calculated using UCINET software, which produces a series of alternative measures (for any categorical variable) for each person in the network. “Yules Q” is a measure of similarity with ranges from −1 for perfect heterophily to +1 for perfect homophily. A value 0 means no pattern of homophily. “Correlation” calculates the correlation between the presence or absence of a tie between each ego and each alter in the network and a vector indicating ego and alter similarity on the selected attribute (gender); interpretation is the same as for Yules Q.

## RESULTS

From 50 invitations submitted, 49 valid questionnaires were obtained corresponding to the same number of participants. Every participant reported having contact with patients with diabetes as part of their work. Participants were from primary care (31, 63%) and secondary care (18, 37%). General practitioners and practice nurses were located in 17 different general practices, while secondary care was treated as a single centre (located in a hospital). General practitioners and specialist nurses were the two groups most represented in this study. Overall, 32 participants were female and 17 male, the numbers in primary care were similar but the majority of the secondary care participants were female. Table 1 reports the distribution of health professionals who participated in the study by profession.

**Table 1:**
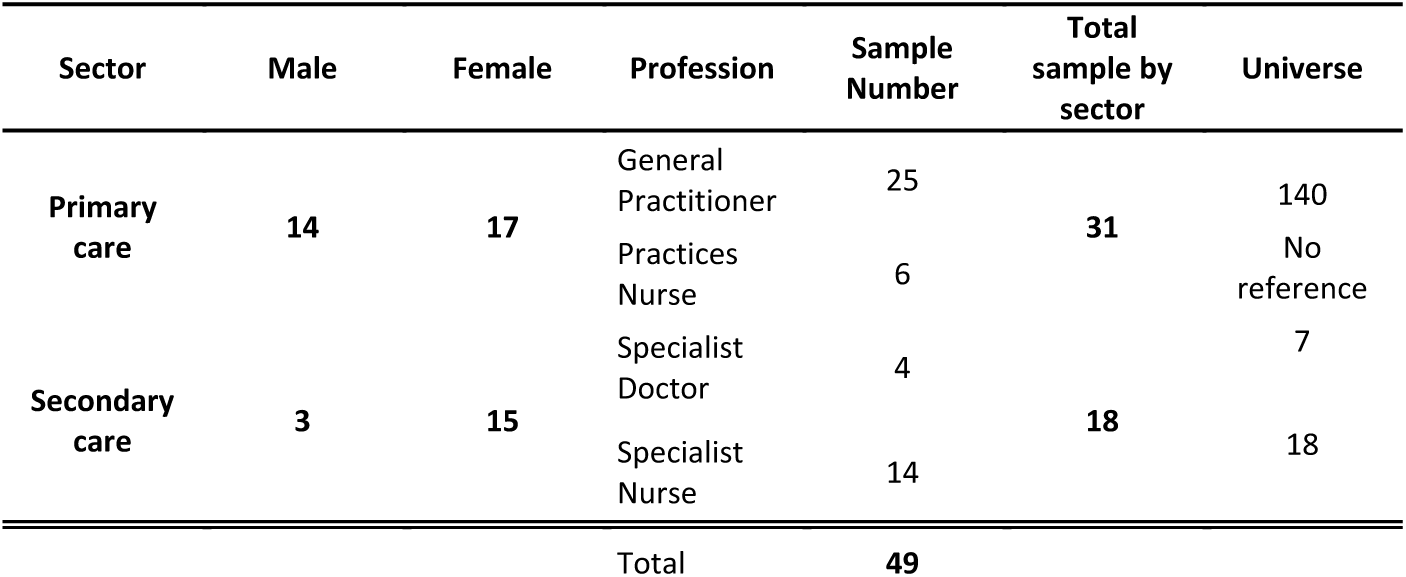
Participant distribution by sector, gender and profession.

### Results of the Corrected Metric Homophily

Table 2 shows that it was possible to calculate gender-homophily in 40 (82%) actors (9 actors did not have interactions within the network). There were 6 actors with rank 0 that mean they do not have any gender preference, 2 actors with a negative rank that represent actors who prefer to interact with people of the opposite gender and 32 actors with a positive rank who like to interact with other actors of the same gender.

**Table 2.** Network Metric Homophily

| Participant | Gender | Ties | Ties opposite sex | Same sex could cited but did | Different sex could cited | Rank homophily |
| --- | --- | --- | --- | --- | --- | --- |
|  |  | same sex |  | not | but did not |  |
| SCN010 | 0 | 16 | 11 | 13 | 7 | -0.07 |
| GD00332 | 1 | 2 | 3 | 16 | 26 | -0.03 |
| GD00114 | 1 | 1 | 1 | 17 | 28 | 0.00 |
| GD00312 | 0 | 1 | 1 | 28 | 17 | 0.00 |
| GD00535 | 1 | 1 | 1 | 17 | 28 | 0.00 |
| GD00930 | 0 | 1 | 1 | 28 | 17 | 0.00 |
| GN00212 | 0 | 1 | 1 | 28 | 17 | 0.00 |
| GN00628 | 0 | 2 | 2 | 27 | 16 | 0.00 |
| GD00926 | 0 | 2 | 1 | 27 | 17 | 0.04 |
| GD00832 | 1 | 1 | 2 | 17 | 27 | 0.05 |
| SCS003 | 1 | 11 | 19 | 7 | 10 | 0.07 |
| SCN001 | 0 | 16 | 9 | 13 | 9 | 0.08 |
| SCS009 | 1 | 10 | 18 | 8 | 11 | 0.09 |
| GN00632 | 0 | 1 | 2 | 28 | 16 | 0.10 |
| GN00832 | 0 | 1 | 2 | 28 | 16 | 0.10 |
| SCN013 | 0 | 1 | 2 | 28 | 16 | 0.10 |
| GD00330 | 0 | 1 | 0 | 28 | 18 | 0.12 |
| GN00326 | 0 | 1 | 0 | 28 | 18 | 0.12 |
| GD00324 | 1 | 0 | 1 | 18 | 28 | 0.14 |
| GD00724 | 0 | 0 | 1 | 29 | 17 | 0.14 |
| GD00628 | 1 | 2 | 6 | 16 | 23 | 0.16 |
| GD00723 | 1 | 1 | 0 | 17 | 29 | 0.19 |
| SCN004 | 0 | 13 | 5 | 16 | 13 | 0.19 |
| GD00512 | 1 | 0 | 2 | 18 | 27 | 0.20 |
| GN00514 | 0 | 0 | 2 | 29 | 16 | 0.20 |
| SCN007 | 0 | 0 | 2 | 29 | 16 | 0.20 |
| GD00629 | 0 | 3 | 0 | 26 | 18 | 0.21 |
| SCN009 | 0 | 3 | 0 | 26 | 18 | 0.21 |
| GD00335 | 1 | 1 | 5 | 17 | 24 | 0.21 |
| GD00514 | 1 | 1 | 5 | 17 | 24 | 0.21 |
| SCC005 | 0 | 14 | 5 | 15 | 13 | 0.22 |
| SCN011 | 0 | 18 | 7 | 11 | 11 | 0.23 |
| SCN012 | 0 | 13 | 4 | 16 | 14 | 0.24 |
| GD00310 | 1 | 0 | 3 | 18 | 26 | 0.24 |
| SCN014 | 0 | 12 | 3 | 17 | 15 | 0.27 |
| SCN002 | 0 | 13 | 3 | 16 | 15 | 0.29 |
| SCN003 | 0 | 13 | 3 | 16 | 15 | 0.29 |
| SCN008 | 0 | 13 | 3 | 16 | 15 | 0.29 |
| SCS011 | 0 | 13 | 3 | 16 | 15 | 0.29 |
| SCS010 | 1 | 2 | 13 | 16 | 16 | 0.35 |

The homophily distribution for the entire network is presented in figure 1. Most values of the homophily near 0, suggesting there is no pattern of homophily by gender in this network.

**Figure 1.**
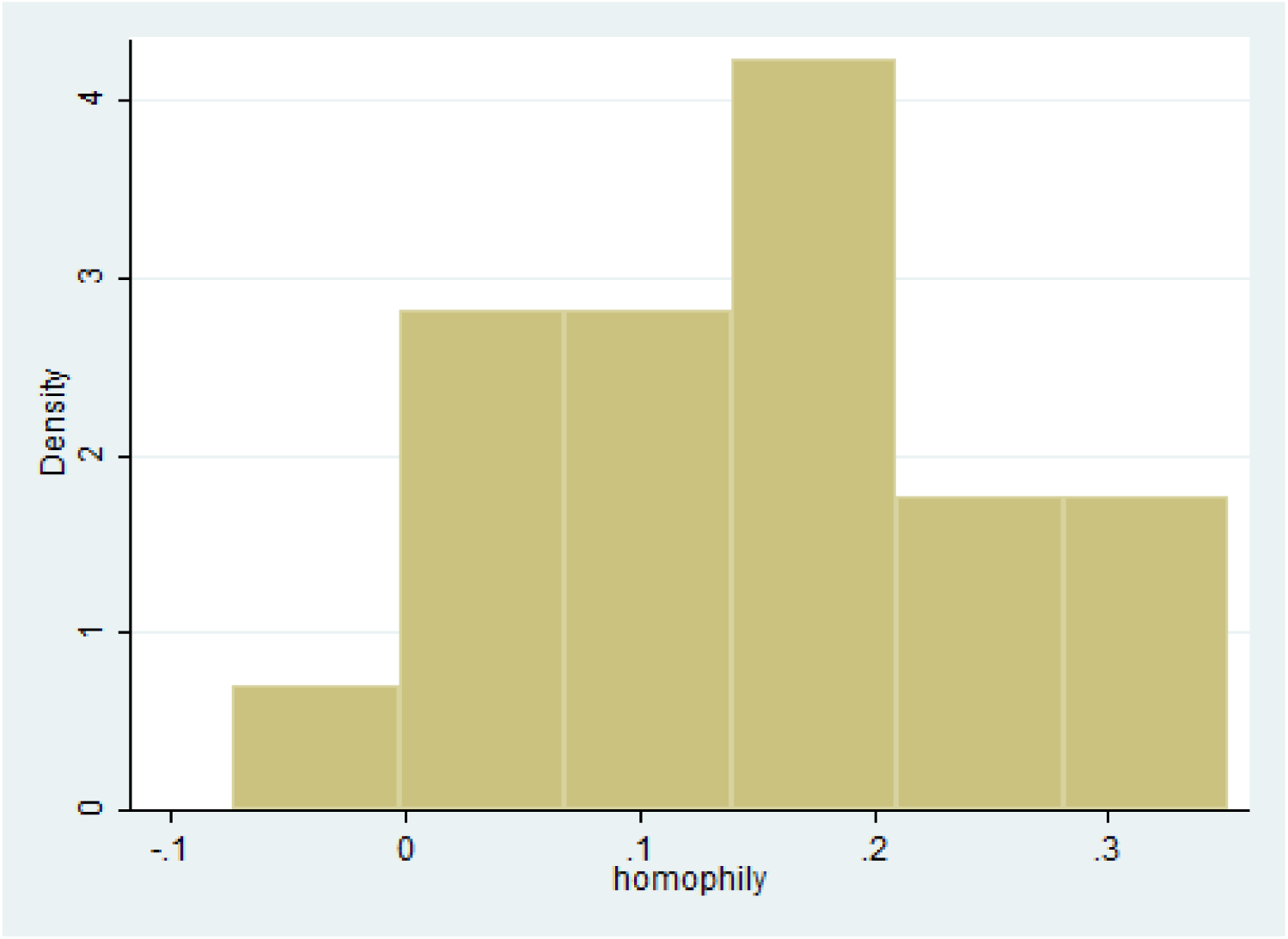
Homophily distribution histogram.

### Ego-net composition

Table 3 shows the homophily ranking using UCINET. The correlation indicator shows values close to 0, suggesting that there are not patterns of homophily by gender in this network.

**Table 3:**
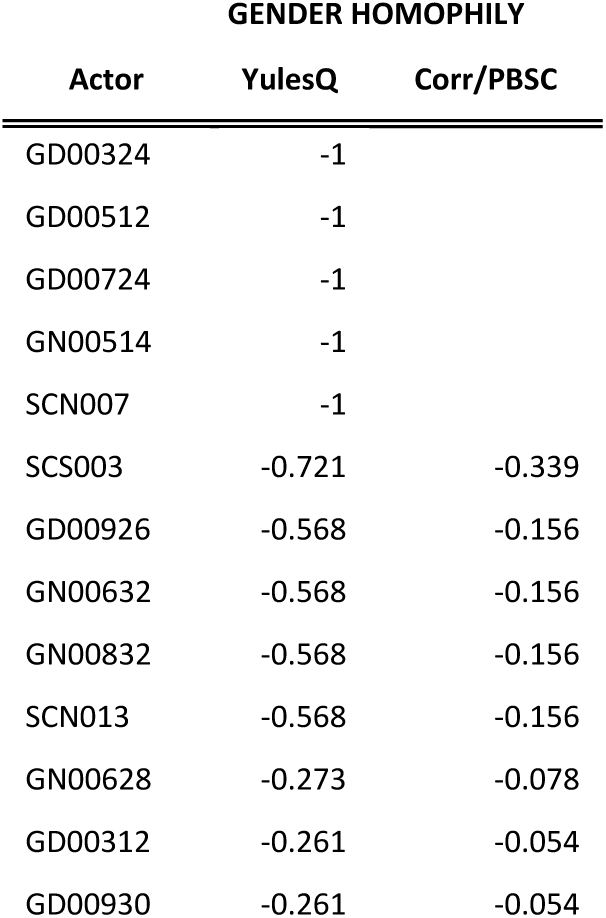
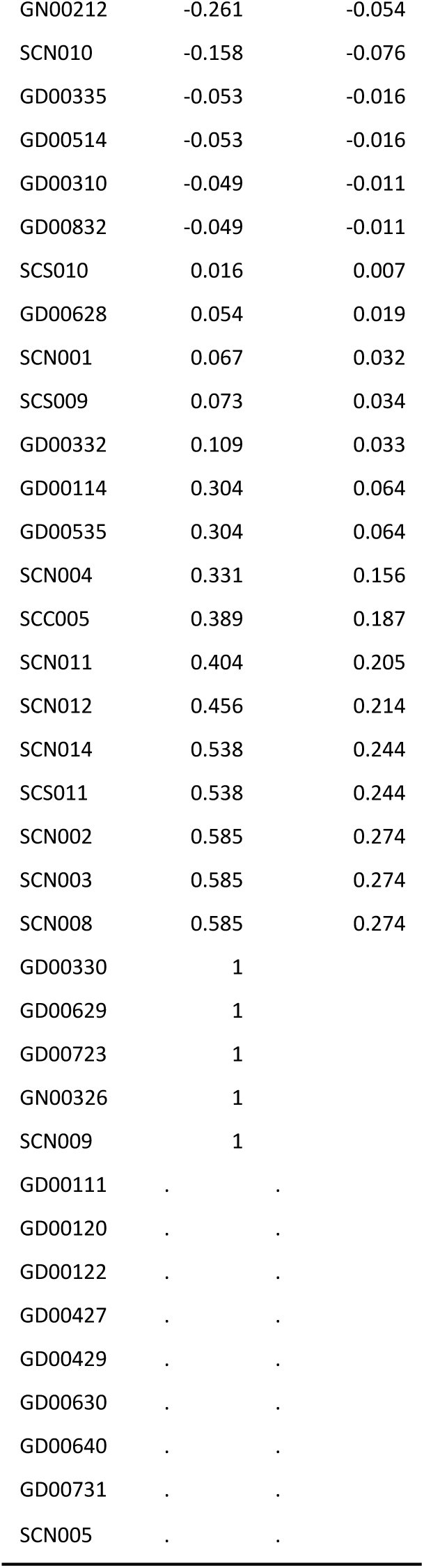
Gender homophily using UCINET

| GENDER HOMOPHILY |  |  |
| --- | --- | --- |
| Actor | YulesQ | Corr/PBSC |
| GD00324 | -1 |  |
| GD00512 | -1 |  |
| GD00724 | -1 |  |
| GN00514 | -1 |  |
| SCN007 | -1 |  |
| SCS003 | -0.721 | -0.339 |
| GD00926 | -0.568 | -0.156 |
| GN00632 | -0.568 | -0.156 |
| GN00832 | -0.568 | -0.156 |
| SCN013 | -0.568 | -0.156 |
| GN00628 | -0.273 | -0.078 |
| GD00312 | -0.261 | -0.054 |
| GD00930 | -0.261 | -0.054 |
| GN00212 | -0.261 | -0.054 |
| SCN010 | -0.158 | -0.076 |
| GD00335 | -0.053 | -0.016 |
| GD00514 | -0.053 | -0.016 |
| GD00310 | -0.049 | -0.011 |
| GD00832 | -0.049 | -0.011 |
| SCS010 | 0.016 | 0.007 |
| GD00628 | 0.054 | 0.019 |
| SCN001 | 0.067 | 0.032 |
| SCS009 | 0.073 | 0.034 |
| GD00332 | 0.109 | 0.033 |
| GD00114 | 0.304 | 0.064 |
| GD00535 | 0.304 | 0.064 |
| SCN004 | 0.331 | 0.156 |
| SCC005 | 0.389 | 0.187 |
| SCN011 | 0.404 | 0.205 |
| SCN012 | 0.456 | 0.214 |
| SCN014 | 0.538 | 0.244 |
| SCS011 | 0.538 | 0.244 |
| SCN002 | 0.585 | 0.274 |
| SCN003 | 0.585 | 0.274 |
| SCN008 | 0.585 | 0.274 |
| GD00330 | 1 |  |
| GD00629 | 1 |  |
| GD00723 | 1 |  |
| GN00326 | 1 |  |
| SCN009 | 1 |  |
| GD00111 | . | . |
| GD00120 | . | . |
| GD00122 | . | . |
| GD00427 | . | . |
| GD00429 | . | . |
| GD00630 | . | . |
| GD00640 | . | . |
| GD00731 | . | . |
| SCN005 | . | . |

#### Density of interaction by gender

Density of interactions was measured by women and men in this network. As a shown figure 2, there are denser interactions between women than between men.

**Figure 2.**
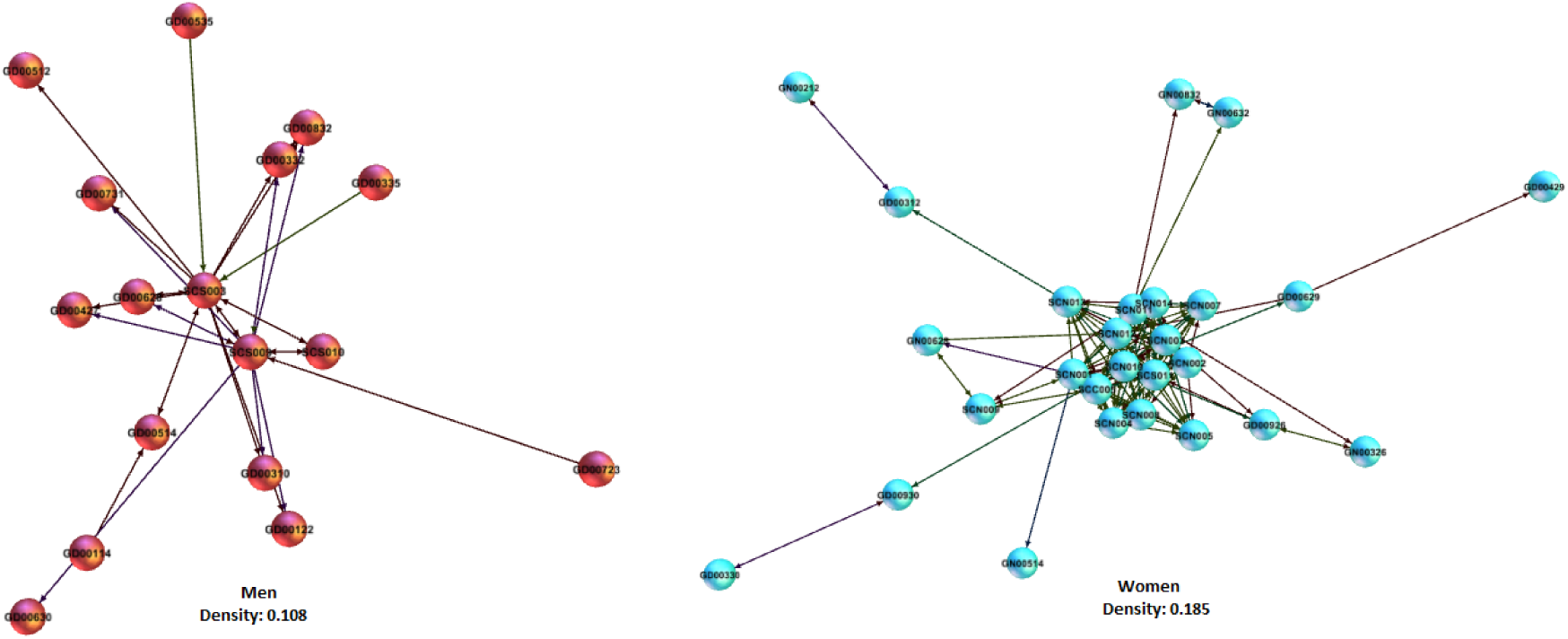
Map of the interactions by gender. Whole network density: 0.156

## DISCUSSION

Using two different methods we found there was no gender homophily in the patterns of interaction between health professionals at the primary and secondary care interface in the diabetes service in Auckland New Zealand. This study confirms strong relationships and tie formation with other colleagues performing collaborative behaviours in similar specialties and interacting organizations, suggesting that gender homophily is not one of the main factors affecting network development in this service. A second aspect to highlight is that women established more interactions with other women particularly within secondary care, in contrast with men. There is an important implication of this pattern, in terms of the speed for spread knowledge and the effectiveness to reach actors in the periphery of the network.

Limitations of this study include the cross-sectional design which does not allow identification of causal relationships. The sample was relatively small and was derived from volunteers within a single primary health organisation and referral hospital, and may not represent all health professionals among the diabetes care programme.

### Conclusion

This study in diabetes managed care network found little evidence to support the impact of gender homophily on communication exchange. This contrasts with other studies in the health care context. Existing literature provides a good understanding of how gender affect network development, but networks are dynamic and increase and decay over the time. This is the reason why research on social networks requires studying interactions within individuals over the time. It will be necessary to identify possible associations between gender and other set of characteristics such as profession and education background, in order to identify potential overlaps and interactions between them that may explain patterns of homophily in healthcare context.

## Data Availability

No data is available

## Authors’ contributions

NA Collected and analysed the data. MM, TK and PC interpreted the data. All authors conceived and designed the study, and drafted the manuscript.

## Acknowledge

The authors would like to thank PROCARE PHO and Counties Manukau DHB for their help with selecting the sample.

